# On the increasing role of older adolescents and younger adults during the SARS-CoV-2 epidemic in Mexico

**DOI:** 10.1101/2020.06.10.20127795

**Authors:** Dalia Stern, Martin Lajous, Blanca De la Rosa, Edward Goldstein

## Abstract

**Background:** During the first months of the SARS-CoV-2 pandemic, Mexico implemented a national lockdown followed by post-lockdown mitigation.

**Methods:** We used daily number of SARS-CoV-2-confirmed hospitalizations (by date of symptom onset) to assess the changes in the incidence of individuals between the age of 10-59 years during the epidemic in Mexico. For each age group g, we computed the proportion E(g) of individuals in that age group among all cases aged 10-59y during the early lockdown period (April 20–May 3, 2020), and the corresponding proportion L(g) during the late lockdown period (May 18-31, 2020) and post-lockdown mitigation (June 15-28, 2020). For each later period (late lockdown or post-lockdown), we computed the proportion ratios relative to the early lockdown period PR(g)=L(g)/E(g). For each pair of age groups g1,g2, PR(g1)> PR(g2) is interpreted as a relative increase in SARS-CoV-2 infections in the age group g1 compared to g2 for the late lockdown and post-lockdown periods vs. the early lockdown period.

**Results:** For the late lockdown period, the highest PR estimates belong to persons aged 15-19y (PR=1.69(95%CI(1.05, 2.72))) and 20-24y (PR=1.43(1.10,1.86)). For the post-lockdown period, the highest PR estimates were also in age groups 15-19y (PR=2.05(1.30, 3.24)) and 20-24y (PR=1.49(1.15,1.93)). These estimates were higher in persons 15-24y compared to those ≥30y.

**Conclusions:** Our results suggest that adolescents and younger adults had an increased relative incidence during late lockdown and the post-lockdown mitigation periods. The role of these age groups during the epidemic should be considered when implementing future pandemic response efforts.

## INTRODUCTION

The SARS-CoV-2 epidemic in Mexico is growing, with 1,070,487 cases and 103,597 deaths recorded by November 25^th^, 2020 [1]. In Mexico, on March 23^rd^ (and until May 30^th^) a call for a nationwide lock-down was made [2,3]. On June 1^st^, Mexico implemented post-lockdown mitigation strategies that depending on community transmission and hospital capacity, eased some of the initial restrictions [4]. To understand the transmission of SARS-CoV-2 under different mitigation strategies, it is important to study the role that different age groups have played on propagating the spread of the virus. Variations in transmission of SARS-CoV-2 by age may take place during the course of an epidemic due to changing mixing patterns [5], which in turn has implications for epidemic control.

Evidence has accumulated that susceptibility to SARS-CoV-2 infection increases with age [6, 7]. Yet, this does not suggest that the oldest groups in a population necessarily play the leading role in the spread of SARS-CoV-2 in the community. Actually, several serological studies suggest that adolescents and younger adults often experience the highest cumulative rates of infection [8-15]. In England, the highest rates of SARS-CoV-2 infection during the Fall of 2020 are in persons aged 18-24y and 13-17y (Figure 8 in [16]). Under the physical distancing measures implemented in Mexico, mixing patterns for individuals in different age groups may be quite different compared to regular mixing patterns [5,6]. Also, similar to other low- and middle-income countries, a sizeable proportion of Mexico’s population are relatively young informal workers who live from day to day for whom shelter-in-place policies may represent a significant economic burden. It is unknown what role do younger adults/older adolescents and other age groups play in propagating the SARS-CoV-2 epidemic in Mexico. To address this question, we estimate temporal changes in SARS-CoV-2 incidence by age group during and after the national lockdown period. We applied the methodology developed previously [17-19] to assess the temporal changes in the incidence of different age groups of individuals between the age of 10-59 years during the epidemic in Mexico.

## METHODS

### Data sources

Information on daily hospitalized COVID-19 cases by age group was obtained from the *Dirección General de Epidemiología* in Mexico. We retrieved data on reported hospitalized cases with PCR-confirmed SARS-CoV-2 infection with available information on the date of symptom onset on September 28, 2020. We excluded healthcare workers because of the significant non-community transmission in that population group. We also excluded cases from private hospitals because it is unclear how cases were ascertained and whether case-ascertainment has changed over time. We also excluded non-hospitalized cases because testing for non-severe COVID-19 may have changed over time for different age groups, while the criteria for testing of cases requiring hospitalization for SARS-CoV-2 infection have been consistent over time.

### Relative change in SARS-CoV-2 infection by age-group

We included laboratory confirmed hospitalizations in ten 5-year age groups: 10-14 years through 55-59 years. Older adults were not included because of potential temporal changes in ascertainment, as well as presence of some hospitalizations stemming from infections that do not reflect community transmission (e.g., infections in long-term care facilities, with rates of infection in those facilities being quite higher than in the corresponding age groups in the community). We excluded children aged under 10 years for two reasons. First, ascertainment of infection in those age groups might have changed with time as more severe episodes in younger children appeared as the epidemic progressed. Second, there is evidence of lower susceptibility to infection for children aged under 10 years compared to adults and older adolescents [20], and those children are unlikely to play a significant role in the progression of the epidemic.

We selected three periods: April 20 – May 3 (early lockdown period, we selected this date because earlier numbers of cases in certain age groups were limited and may have rendered our estimates unstable), 18-31 May (late lockdown period) and 15–25 June (post-lockdown mitigation, starting two weeks after the national lockdown to detect changes in symptom onset).

We applied a previously described procedure [17-19] to estimate the age-specific proportion ratios for late lockdown (May 18-31, 2020) and post-lockdown mitigation (June 15-28, 2020) relative to the early lockdown (April 20–May 3, 2020) as follows: let *E(g)* be the number of hospitalization with confirmed SARS-CoV-2 infection in age group *g* andΣ_*h*_ *E*(*h*) the total number of cases in age groups h=1 (10–14 years) to h =10 (55–59 years) during early lockdown, and *L(g)* and Σ_*h*_ *E*(*h*) be the corresponding numbers during late lockdown (and post-lockdown). The proportion ratio (PR) statistic in age group *g* for the late vs. early lockdown comparison is

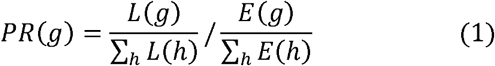

that is, the ratio between the proportion of cases in age group *g* among all cases in the late lockdown period and the proportion of cases in age group g in the early lockdown. The logarithm ln(*PR*(*g*)) of the PR(*g)* is approximately normally distributed [21] with the standard error:

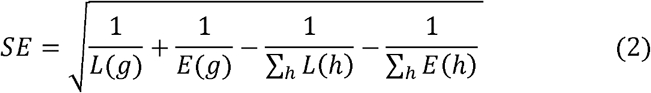

We repeated these calculations after replacing the cases in the late lockdown by the cases in the post-lockdown period.

To examine whether the PR in certain age groups are significantly higher than in others, we consider the corresponding pairwise odds ratios (ORs). For each pair of age groups *g*1 and *g*2, the proportion ratios *PR*(*g*1) and *PR*(*g*2) are compared using the odds ratio (OR)

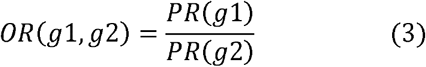

It follows from equation 1 that *OR*(*g*1,*g*2) equals

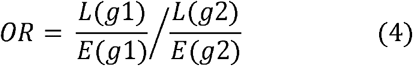

which is the OR for a hospitalized case with confirmed SARS-CoV-2 infection to be in age group *g*1 vs *g*2 for the early lockdown period and the late lockdown period. We repeated these calculations for the comparison of early lockdown to post-lockdown. Estimates for pairwise OR were performed using Fisher’s exact test.

## RESULTS

Table 1 shows the number of hospitalizations with confirmed SARS-CoV-2 infection in the different age groups (10-14 through 55-59 years) for the early lockdown (April 20–May 3, 2020), the late lockdown (May 18-31, 2020) and the post-lockdown (June 15-28, 2020) periods, as well as the corresponding estimates of the PR statistic (equations 1,2).

**Table 1.** Number of hospitalizations with conformed SARS-CoV-2 infection by age groups (10-14 through 55-59 years) and time period, and the estimates of the prevalence ratio (PR) statistic (n=23,013)

|  | Early lockdown period |  | Late lockdown period |  | Post-lockdown period |  | Prevalence Ratio<br>PR (95% CI) |  |
| --- | --- | --- | --- | --- | --- | --- | --- | --- |
| Age Group, y | April 20-26 | April 27 - May 3 | May 18-24 | May 25-31 | June 15-21 | June 22-28 | Late vs. early lockdown period | Post-lockdown vs. early lockdown period |
| 10–14 | 9 | 7 | 13 | 16 | 28 | 21 | 1.13 (0.62, 2.09) | 1.66 (0.94, 2.91) |
| 15–19 | 11 | 12 | 35 | 27 | 45 | 42 | **1.69 (1.05, 2.72)** | **2.05 (1.30, 3.24)** |
| 20–24 | 34 | 44 | 87 | 91 | 115 | 100 | **1.43 (1.10, 1.86)** | **1.49 (1.15, 1.93)** |
| 25–29 | 103 | 108 | 195 | 179 | 216 | 199 | 1.11 (0.94, 1.31) | 1.07 (0.91, 1.25) |
| 30–34 | 164 | 186 | 288 | 276 | 347 | 355 | 1.01 (0.89, 1.15) | 1.09 (0.96, 1.23) |
| 35–39 | 251 | 239 | 385 | 430 | 461 | 454 | 1.04 (0.94, 1.16) | 1.01 (0.91, 1.12) |
| 40–44 | 330 | 385 | 559 | 555 | 620 | 587 | 0.98 (0.89, 1.06) | 0.91 (0.84, 1.00) |
| 45–49 | 465 | 547 | 741 | 747 | 869 | 878 | 0.92 (0.86, 0.99) | 0.93 (0.87, 1.00) |
| 50–54 | 555 | 581 | 865 | 902 | 1,030 | 957 | 0.97 (0.91, 1.04) | 0.95 (0.89, 1.01) |
| 55–59 | 544 | 603 | 915 | 968 | 1,114 | 1,123 | 1.03 (0.96, 1.10) | 1.06 (0.99, 1.12) |
| Total | 2466 | 2712 | 4083 | 4191 | 4845 | 4716 |  |  |

For the late lockdown period vs. the early lockdown, the highest PR (95% CI) estimates belong to persons aged 15-19 years (PR=1.69(1.05, 2.72)) and 20-24 years (PR=1.43(1.10,1.86)) (Table 1). The PR estimates in persons aged over 30 years were significantly lower compared to persons aged 15-24 years. Table 2 gives the estimates of ORs for different pairs of age groups (10-14y through 55-59y) for a hospitalized case with confirmed SARS-CoV-2 infection to be during the period May 18-31 vs. April 20-May 3 (equation 4). Our results suggest that for persons aged 15-24y, the corresponding OR relative to any age group over 30y is above 1.

**Table 2.** Odds ratios for different pairs of age groups for a hospitalized case with confirmed SARS-CoV-2 infection to occur between the late lockdown (May 18-31) vs. the early lockdown period (April 20-May 3)

|  | 15-19 | 20-24 | 25-29 | 30-34 | 35-39 | 40-44 | 45-49 | 50-54 | 55-59 |
| --- | --- | --- | --- | --- | --- | --- | --- | --- | --- |
| 10-14 | 0.67<br>(0.29, 1.58) | 0.79<br>(0.39, 1.66) | 1.02<br>(0.52, 2.06) | 1.12<br>(0.58, 2.25) | 1.09<br>(0.57, 2.17) | 1.16<br>(0.61, 2.31) | 1.23<br>(0.64, 2.44) | 1.17<br>(0.61, 2.31) | 1.10<br>(0.58, 2.19) |
| 15-19 |  | 1.18<br>(0.67, 2.15) | 1.52<br>(0.90, 2.65) | 1.67<br>(1.00, 2.88) | 1.62<br>(0.97, 2.78) | **1.73**<br>**(1.05, 2.95)** | **1.83**<br>**(1.11, 3.12)** | **1.73**<br>**(1.05, 2.95)** | 1.64<br>(1.00, 2.79) |
| 20-24 |  |  | 1.29<br>(0.93, 1.79) | **1.42**<br>**(1.04, 1.93)** | **1.37**<br>**(1.02, 1.86)** | **1.46**<br>**(1.10, 1.97)** | **1.55**<br>**(1.17, 2.08)** | **1.47**<br>**(1.11, 1.96)** | **1.39**<br>**(1.05, 1.86)** |
| 25-29 |  |  |  | 1.10<br>(0.88, 1.37) | 1.07<br>(0.87, 1.31) | 1.14<br>(0.93, 1.39) | 1.21<br>(1.00, 1.46) | 1.14<br>(0.94, 1.38) | 1.08<br>(0.89, 1.30) |
| 30-34 |  |  |  |  | 0.97<br>(0.81, 1.16) | 1.03<br>(0.88, 1.22) | 1.10<br>(0.94, 1.28) | 1.04<br>(0.89, 1.21) | 0.98<br>(0.84, 1.14) |
| 35-39 |  |  |  |  |  | 1.07<br>(0.92, 1.24) | 1.13<br>(0.98, 1.30) | 1.07<br>(0.93, 1.23) | 1.01<br>(0.88, 1.16) |
| 40-44 |  |  |  |  |  |  | 1.06<br>(0.93, 1.20) | 1.00<br>(0.89, 1.13) | 0.95<br>(0.84, 1.07) |
| 45-49 |  |  |  |  |  |  |  | 0.95<br>(0.85, 1.06) | 0.90<br>(0.80, 1.00) |
| 50-54 |  |  |  |  |  |  |  |  | 0.95<br>(0.85, 1.05) |

Odds ratios (ORs) for different pairs of age groups (10-14y through 55-59y) for a hospitalized case with confirmed SARS-CoV-2 infection in Mexico to be during the period of May 18-31 vs. April 20 – May 3 (equation 4)

For the post-lockdown period vs. the early lockdown, the highest PR (95% CI) estimates belong to persons aged 15-19 years (PR=2.05(1.30, 3.24)) and 20-24 years (PR=1.49(1.15,1.93)) (Table 1). The PR estimates in persons aged over 30 years were significantly lower compared to persons aged 15-24 years. Table 3 shows the estimates of ORs during the post-lockdown period of June 15-28 vs. the lockdown period April 20-May 3. The corresponding OR for persons aged 15-24y relative to any age group over 30y is significantly above 1 for the post-lockdown period compared to the early lockdown period. This suggests a relative increase in the incidence of SARS-CoV-2 infection in persons aged 15-24y compared to persons aged 30-59y for the post-lockdown period relative to the early lockdown period.

**Table 3.** Odds ratios for different pairs of age groups for a hospitalized case with confirmed SARS-CoV-2 infection to occur between the post-lockdown (June 15-18) vs. the early lockdown period (April 20-May 3)

|  | 15-19 | 20-24 | 25-29 | 30-34 | 35-39 | 40-44 | 45-49 | 50-54 | 55-59 |
| --- | --- | --- | --- | --- | --- | --- | --- | --- | --- |
| 10-14 | 0.81<br>(0.37, 1.80) | 1.11<br>(0.58, 2.19) | 1.56<br>(0.85, 3.00) | 1.53<br>(0.84, 2.92) | 1.64<br>(0.91, 3.12) | 1.81<br>(1.00, 3.44) | 1.77<br>(0.99, 3.36) | 1.75<br>(0.97, 3.31) | 1.57<br>(0.87, 2.97) |
| 15-19 |  | 1.37<br>(0.79, 2.44) | **1.92**<br>**(1.16, 3.28)** | **1.89**<br>**(1.16, 3.18)** | **2.03**<br>**(1.25, 3.40)** | **2.24**<br>**(1.39, 3.75)** | **2.19**<br>**(1.36, 3.66)** | **2.16**<br>**(1.34, 3.61)** | **1.94**<br>**(1.21, 3.24)** |
| 20-24 |  |  | **1.40**<br>**(1.02, 1.93)** | **1.37**<br>**(1.02, 1.86)** | **1.48**<br>**(1.11, 1.98)** | **1.63**<br>**(1.23, 2.18)** | **1.60**<br>**(1.21, 2.12)** | **1.58**<br>**(1.20, 2.09)** | **1.41**<br>**(1.07, 1.87)** |
| 25-29 |  |  |  | 0.98<br>(0.79, 1.22) | 1.05<br>(0.86, 1.29) | 1.17<br>(0.96, 1.42) | 1.14<br>(0.94, 1.37) | 1.12<br>(0.93, 1.35) | 1.01<br>(0.84, 1.21) |
| 30-34 |  |  |  |  | 1.07<br>(0.90, 1.28) | **1.19**<br>**(1.01, 1.40)** | 1.16<br>(1.00, 1.35) | 1.15<br>(0.99, 1.33) | 1.03<br>(0.89, 1.19) |
| 35-39 |  |  |  |  |  | 1.11<br>(0.96, 1.28) | 1.08<br>(0.94, 1.24) | 1.07<br>(0.93, 1.22) | 0.96<br>(0.84, 1.09) |
| 40-44 |  |  |  |  |  |  | 0.98<br>(0.87, 1.11) | 0.97<br>(0.86, 1.09) | **0.87**<br>**(0.77, 0.97)** |
| 45-49 |  |  |  |  |  |  |  | 0.99<br>(0.89, 1.10) | **0.89**<br>**(0.80, 0.98)** |
| 50-54 |  |  |  |  |  |  |  |  | **0.90**<br>**(0.81, 0.99)** |

Odds ratios (ORs) for different pairs of age groups (10-14y through 55-59y) for a hospitalized case with confirmed SARS-CoV-2 infection in Mexico to be during the period of June 15-28 vs. April 20 – May 3 (equation 4)

## DISCUSSION

We applied the previously developed methodology [17-19] to study changes in the relative incidence of SARS-CoV-2 infections in different age groups during national lockdown and post-lockdown mitigation in Mexico. Compared with early lockdown, the greatest relative increase in the incidence of infection belongs to persons aged 15-24 years in both, the late lockdown and post-lockdown mitigation. We note that in England, the highest rates of SARS-CoV-2 infection during the Fall of 2020 are in persons aged 18-24y and 13-17y (Figure 8 in [16]), which is consistent with our findings about the epidemiological importance of older adolescents and younger adults during the epidemic.

We hypothesize there are two potential explanations for the increase in the incidence of SARS-CoV-2 infection in older adolescents and younger adults in Mexico: (i) the need for young informal workers to return to work due to increasing economic burden of physical distancing measures; (ii) fatigue related to adherence to the physical distancing measures, resulting in progressively lesser adherence. Other explanations, such as changing social responsibilities and increased use of public transportation may also apply, and further work is needed to understand those issues to better inform future mitigation efforts. A serological evaluation is underway which may confirm our observations.

Our results are consistent with observations in Germany, where using the same methodology, a higher relative incidence of infection in older adolescents and younger adults following the introduction of physical distancing measures was found [17]. In Spain, using the same methodology as in [17], it was shown that during the initial lockdown period, when non-essential work was allowed, individuals aged 40-64 years had a higher relative incidence of infection compared with the pre-lockdown period. However, during the later strengthened lockdown, older adolescents and younger adults had an increased relative incidence in SARS-CoV-2 infection [22]. Our results, together with these observations, highlight the fact that control measures during a pandemic have differential effectiveness in different age groups.

An important strength of our manuscript was the use of limited data obtained mostly from detected cases to examine the role of different population groups in propagating the spread of infection. However, our study is not without limitations. Our findings could be affected by age-differential changes in case ascertainment, over time or across regions. However, we restricted the analysis to hospitalized cases rather than all confirmed COVID-19 cases in the community because changes in healthcare seeking behavior (e.g., for ambulatory visits), and changes in testing in the outpatient setting might affect the relation between the incidence of SARS-CoV-2 infection and the rates of detected COVID-19 cases. However, such temporal changes are less likely for hospitalized cases, with uniform guidelines for testing hospitalized cases applied in Mexico, and low likelihood of mild cases resulting in hospitalization during certain time periods. Second, the perception of the potential severity of the disease may have changed over time and clinicians, even with an existing case definition, may have preferentially tested certain age groups. We explored whether testing for hospitalized patients in all age groups changed over time and did not find evidence for this (Supplemental Figure). Third, we used date of symptom onset to temporally classify cases from an administrative database. Thus the possibility of error in registration is present. However, this error is probably random and unlikely to affect results. Alternatively, there might be differences across age groups in their recall of the date of onset, yet this seems unlikely. Fourth, it is important to notice that the database from the Ministry of Health it is not updated over time. In other words, we do not know if a case that was originally registered as an ambulatory case was later hospitalized.

In conclusion, our paper provides evidence for an increased relative incidence of SARS-CoV-2 infection among individuals aged 15-24 years when the lockdown interventions were lifted. Our results suggest that the age structure was an important factor in the effect of lockdown interventions. Multigenerational household arrangements are common in Mexico. Therefore, efforts aimed at spreading risk awareness in adolescents and young adults and limiting social interactions for members of certain age groups in certain venues may be considered to stem the increase of COVID-19 incidence in the community in Mexico.

## Data Availability

We used data from Mexico's Ministry of Health available through a collaborative agreement.

## Acknowledgements

We thank Tonatiuh Barrientos and Hector Lamadrid of the Instituto Nacional de Salud Pública and Ruy-López Ridaura of the Mexican Ministry of Health who provided feedback on the results. We also thank all the contributors to the surveillance and investigation of the SARS-CoV-2 epidemic in Mexico. This work was supported by Award Number U54GM088558 from the National Institute of General Medical Sciences (EG)

---

**Supplemental Figure.**
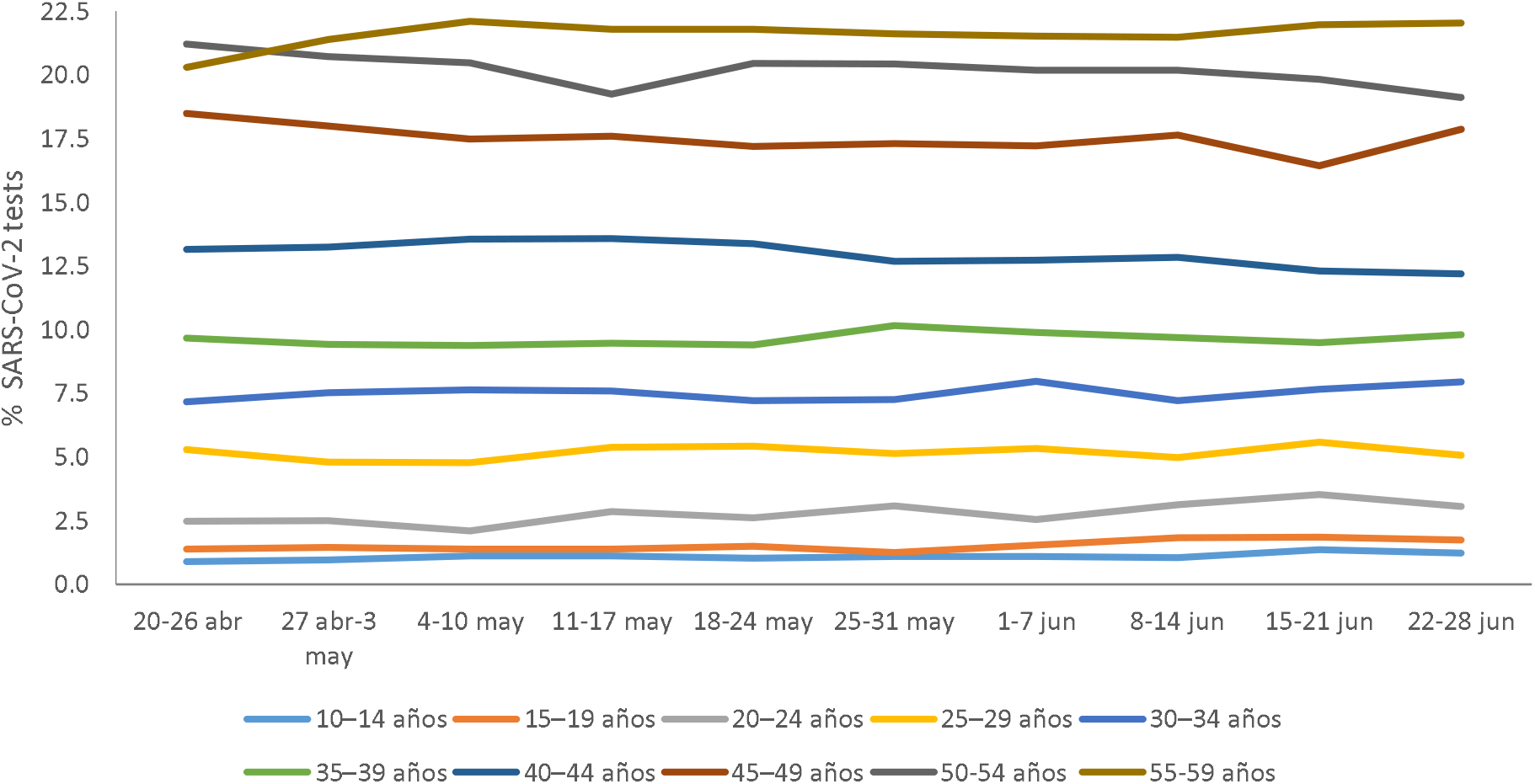
Trends in the distribution of SARS-CoV-2 tests (RT-PCR) by age group among hospitalized individuals.

## Notes

### Competing Interest Statement

The authors have declared no competing interest.

### Funding Statement

None

### Author Declarations

We obtained a waiver for human subjects research review by Instituto Nacional de Salud Publica's IRB. The waiver was signed by Dr. Angelica Angeles Llerenas Chair of the Research Ethics Committee

### Summary of Updates

We used non-public data that allowed for the exclusion of healthcare workers because they may not represent community transmission. We also added a third study period post lockdown mitigation (June 15 to 28, 2020).

